# SARS-CoV-2 titers in wastewater are higher than expected from clinically confirmed cases

**DOI:** 10.1101/2020.04.05.20051540

**Authors:** FQ Wu, A Xiao, JB Zhang, XQ Gu, WL Lee, K Kauffman, WP Hanage, M Matus, N Ghaeli, N Endo, C Duvallet, K Moniz, TB Erickson, PR Chai, J Thompson, EJ Alm

**Affiliations:** Center for Microbiome Informatics and Therapeutics, Departments of Biological Engineering and Civil & Environmental Engineering, Massachusetts Institute of Technology; Singapore-MIT Alliance for Research and Technology, National University of Singapore; University at Buffalo, The State University of New York; Center for Communicable Disease Dynamics, Department of Epidemiology, Harvard T. H. Chan School of Public Health, Boston; Biobot Analytics, Cambridge MA; Division of Medical Toxicology, Department of Emergency Medicine, Brigham and Women’s Hospital; Singapore Center for Environmental Life Sciences Engineering, Asian School of the Environment, Nanyang Technological University, Singapore

## Abstract

Wastewater surveillance may represent a complementary approach to measure the presence and even prevalence of infectious diseases when the capacity for clinical testing is limited. Moreover, aggregate, population-wide data can help inform modeling efforts. We tested wastewater collected at a major urban treatment facility in Massachusetts and found the presence of SARS-CoV-2 at high titers in the period from March 18 - 25 using RT-qPCR. We then confirmed the identity of the PCR product by direct DNA sequencing. Viral titers observed were significantly higher than expected based on clinically confirmed cases in Massachusetts as of March 25. The reason for the discrepancy is not yet clear, and until further experiments are complete, these data do not necessarily indicate that clinical estimates are incorrect. Our approach is scalable and may be useful in modeling the SARS-CoV-2 pandemic and future outbreaks.

## Introduction

Improved understanding of the presence and prevalence of SARS-CoV-2 at a population level can help government and hospital officials implement appropriate policies to mitigate the exponential spread of COVID-19, and diminish the future strain on healthcare facilities. Despite pandemic spread of SARS-CoV-2 worldwide, broad access to testing in the United States (US) has thus far been severely limited. While it is impractical to test every US resident for SARS-CoV-2, the virus has been found in the stool of confirmed COVID-19 patients *(1)*, making it a promising candidate for wastewater-based epidemiology (WBE). WBE can help detect the presence of pathogens across municipalities and estimate population prevalence without individual testing, and inform public health officials of the efficacy of interventions. The closely related virus SARS-CoV was detected in wastewater from Chinese hospitals during the 2002-2003 SARS pandemic *(2)*, and WBE has been used for the early detection and direct mitigation of disease outbreaks in Israel, Egypt and Sweden *(3*–*7)*. We have previously used this technique to measure and map the use of pharmaceuticals across residential communities *(8)*. Here, we describe an analytical technique to extract and detect genetic material from SARS-CoV-2 using wastewater collected at a treatment facility.

## Results

We collected sewage samples at a major urban wastewater treatment facility in Massachusetts. Samples were transported to our laboratory where we conducted viral inactivation and enrichment, nucleic acid extraction, and RT-qPCR. As negative controls, we used biobanked wastewater samples from the same treatment facility taken before the first US case was documented.

Initial testing with PCR using primers specific for the SARS-CoV-2 *S* gene *(9)* indicated that both samples collected from the treatment facility on March 18 had a positive signal for SARS-CoV-2. We confirmed the signal by identifying a PCR product at ∼150 bp. A no-template control for the PCR assays was negative. Sanger sequencing of the PCR products confirmed a 97-98% identity match to the SARS-CoV-2 *S* gene (Figure 1).

**Figure 1.**
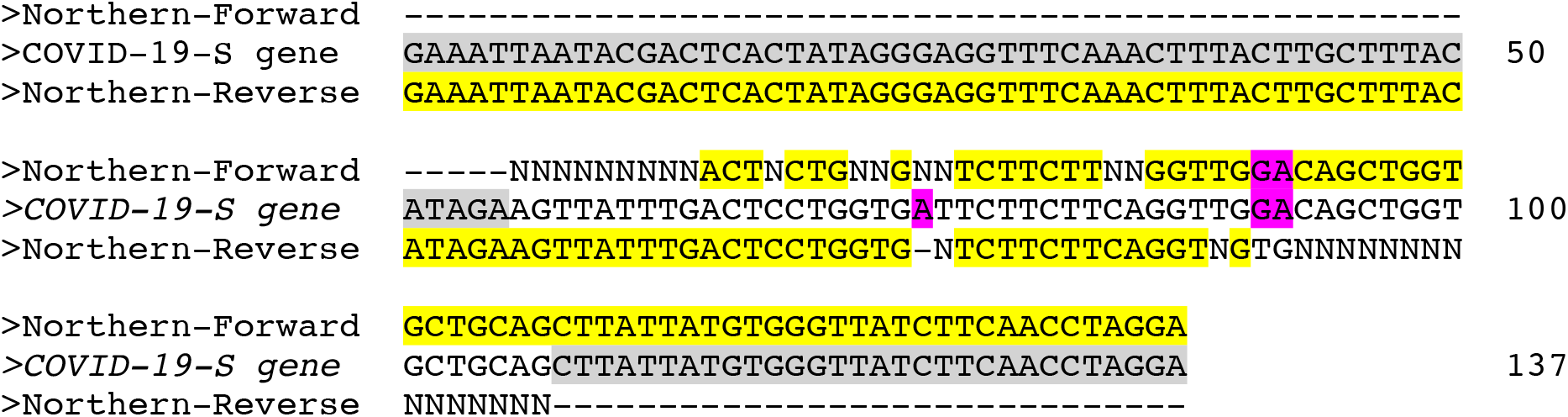
Sanger Sequencing results and alignment to COVID-19 *S* gene. Highlighted grey: forward and reverse primers; highlighted yellow: aligned sequence; highlighted purple: mismatched sequence from either primer. The first 24 bases in the COVID-19 *S* gene is the T7 promoter added by PCR with S-F and S-R primers in Table 1. Results shown only for sequence of template form Northern influent, results from Southern influent were similar.

We next sought to quantify viral titer in sewage, and to establish our viral enrichment protocol using RT-qPCR. We used the US CDC primer/probe sets targeting the N1, N2, and N3 loci of the SARS-CoV-2 nucleocapsid gene to amplify cDNA reverse transcribed from viral RNA (Table 1). We tested different steps in our viral enrichment process. First, we examined raw (unfiltered) sewage precipitated with polyethylene glycol 8000 (PEG), which recovers both bacterial and viral nucleic acids. Next, we looked at samples taken from 0.2 um filtered sewage: we considered both the material collected on the filter and the filtrate. We found the strongest and most consistent results from the PEG-precipitated viral pellet from the 0.2 um filtrate, which was resuspended in Trizol for RNA extraction (Table 2).

**Table 1.**
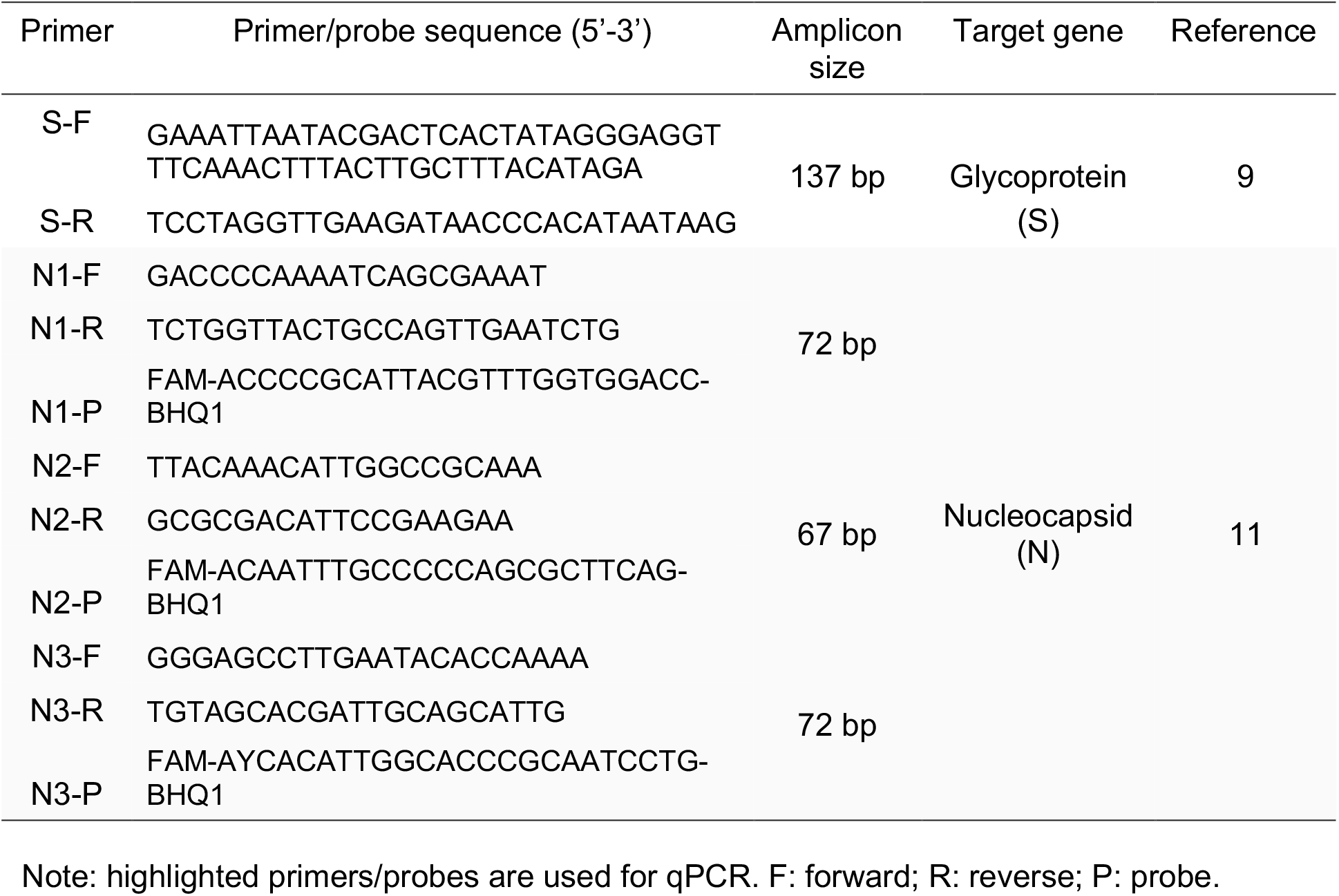
Primers and probes used in this study.

**Table 2.** Mean Ct values for Southern and Northern samples (unfiltered vs filters vs filtrate).

|  | N1 primer | N2 primer | N3 primer |
| --- | --- | --- | --- |
| Southern-Filtrate | 33.87 | 38.39 | 37.12 |
| Southern-Filter | NA | NA | NA |
| Southern-Unfiltered | NA | NA | NA |
| Northern-Filtrate | 34.91 | NA | 37.85 |
| Northern-Filter | 37.86 | NA | 38.14 |
| Northern-Unfiltered | 35.04 | NA | 38.09 |

These results suggest a fairly simple viral enrichment protocol is sufficient to achieve viral identification. Importantly, we included pasteurization (90 min at 60C) as a first step performed before sample containers were opened, increasing the safety of the protocol. Prior work on SARS-CoV-1 indicates that a 30 min heat inactivation at 60C is sufficient to inactivate the virus by over 6 log units *(10)*.

Having shown that it is possible to detect the presence of SARS-CoV-2 in wastewater treatment facility samples, we next looked at whether the viral titer in sewage could be quantified. Standard curves for all three primer sets showed linear behavior over 6 log units (r^2^ ranging from 0.981 to 0.998) using DNA standards (SARS-CoV-2 nucleocapsid gene). We sought to validate our results against other samples, including biobanked samples collected before the first known US case on January 20, which could serve as a relevant negative control. Figure 2 shows estimated viral titer per mL of sewage at the treatment facility over seven different sample dates, with samples from two catchment areas (Southern and Northern) for each date. All four samples from before the first known US SARS-CoV-2 case were negative for virus. In contrast, all 10 samples taken in the range of March 18 - March 25 tested positive (seven out of ten samples hit all the three primers with an average Ct for all samples below 40) for virus by qPCR, using the DNA probes provided by the CDC *(11)*. For each positive sample, we confirmed the presence of a ∼150 bp band after PCR using the S gene primers (data not shown). Thus, our approach is sensitive enough to detect viral titers similar to those currently seen in the treatment facility catchments. Spike-in experiments with purified virus will be needed to establish rigorous limits of detection and are currently underway.

**Figure 2.**
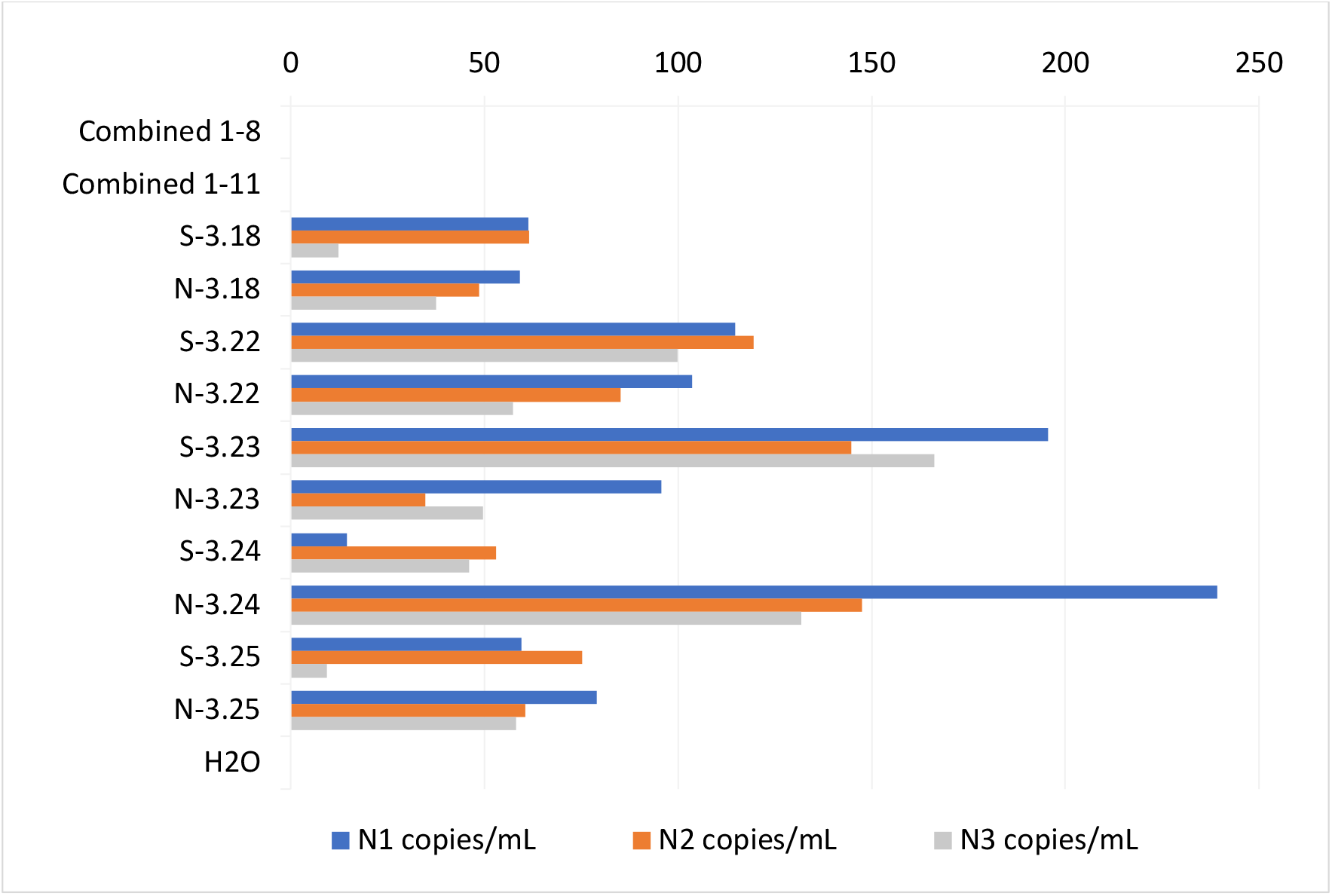
Estimated viral titer per mL of sewage. Estimated gene copies for each of the three CDC primer sets are shown for all dates. Northern (N) and Southern (S) influents are shown separately, except for biobanked January samples, which are combined. H2O represents a PCR negative control.

Finally, we looked at how quickly viral signal degraded during storage at 4C. All samples were received between March 20-25, but samples were stored and reprocessed on April 4. All samples were pasteurized upon initial receipt. Figure 3 shows the variation between measured samples processed on both dates. There was variation between sample runs, but no clear trend toward lower signals at the later timepoint, suggesting samples can be stored for more than a week without significant degradation of signal.

**Figure 3.**
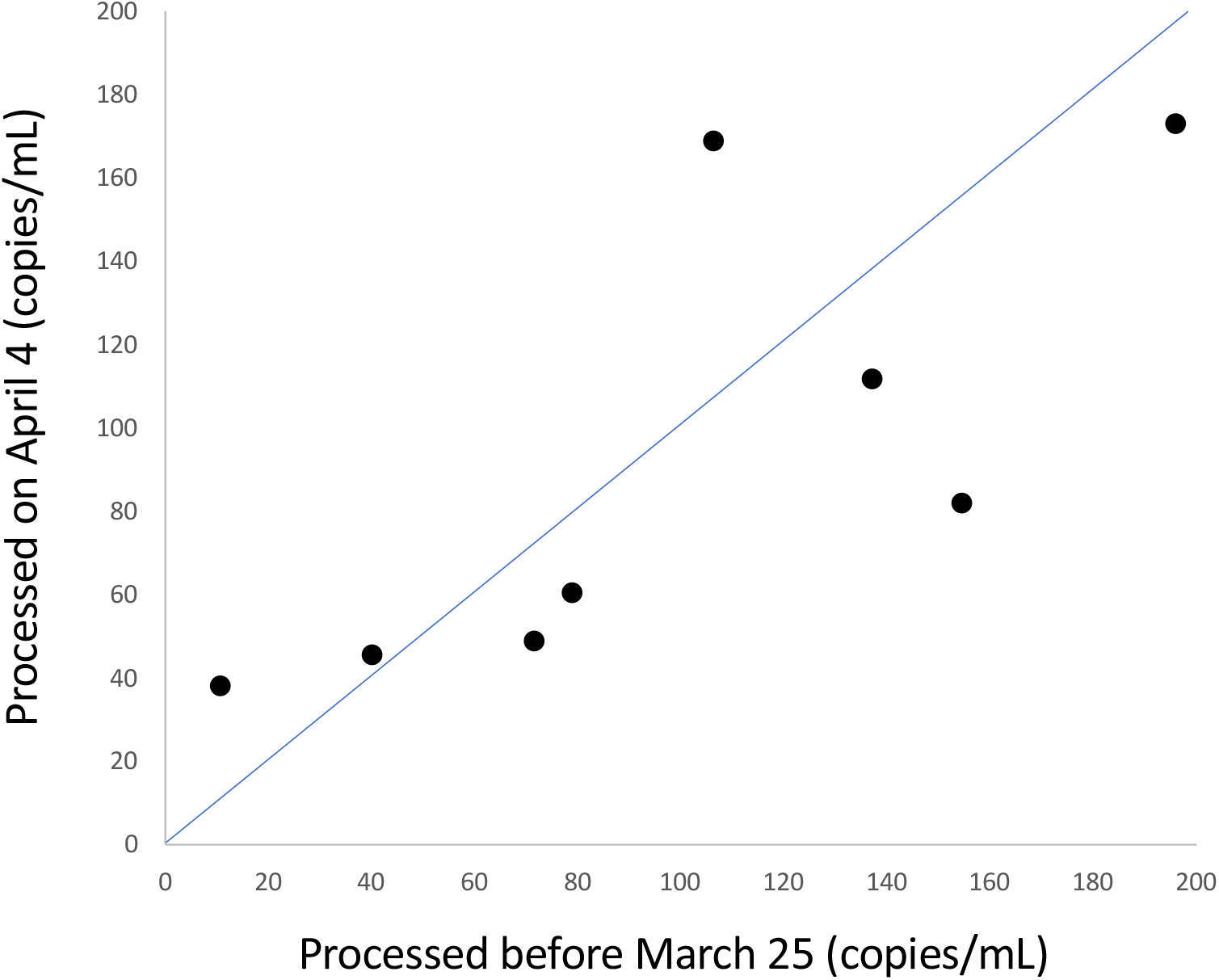
Sample stability over time. Estimated copies per mL sewage estimated from the same samples filtered and processed on different dates. The x-axis shows samples processed between March 20-25, 2020, and are plotted against samples re-processed on April 4, 2020 (samples were pasteurized upon receipt and stored at 4C until processing). The blue line corresponds to x=y. Each point represents an average over all three primer sets for each sample.

## Discussion

These data suggest an order of magnitude estimate of approximately 100 viral particles per mL of sewage, but what conclusions can be drawn about disease prevalence? First, we note that any rigorous conclusions depend on a number of factors that are unknown, and thus additional experiments will be required to calibrate these numbers. Nonetheless, we can estimate an abundance based on the lowest observed values across these samples of ∼10 copies/mL. If we assume typical stool sizes of 200g, diluted into an average volume of 1.36*10^9^ L, and a population of 2.3*10^6^ individuals each producing one stool per day, and we further assume that there is no loss of viral RNA in sewer lines and that excreted viruses are fully suspended in sewage, then we expect the viral titer in feces to be about 3000 times higher than that in sampled raw sewage, or about 30,000 particles per mL.

Estimates of viral load in stool from positive patients are still a matter of uncertainty, but at least one recent publication suggests levels as high as 600,000 viral genomes per mL of fecal material *(12)*. This number would suggest roughly 5% of all fecal samples in the treatment facility catchment were positive for SARS-CoV-2 in the March 18 – 25 period, a number much higher than the 0.026% confirmed for the state of Massachusetts (a similar prevalence is obtained using individual counties represented by the wastewater treatment facility’s catchment or state-wide estimates) on March 25. Another paper reported a maximum observed value close to 30,000,000 viral particles per mL in a single fecal sample *(13)*. If we use this number instead, we would estimate a prevalence of 0.1%, closer to, but still much higher than the number of confirmed clinical cases. Additional data on viral shedding in stool over the course of disease is required to fully interpret these findings.

This discrepancy between confirmed cases and observed viral titers could arise from a number of factors. First, our estimates are based on numbers and assumptions that are currently subject to significant uncertainty, as discussed above, and therefore could be incorrect. However, we note that our calculations are conservative because we assume no loss in viral titer due to degradation, sample processing, or RNA extraction. Second, the estimates of viral titer per positive stool may be too low if a small number of individuals shed very high levels of virus. While this can be tested, it may require extensive testing of individual stools in order to arrive at an accurate estimate. Given the importance of assessing the fraction of SARS-CoV-2 infections that present with symptoms, we note that our results are consistent with the idea that a significant fraction of cases that are not detected with current testing algorithms, and that this fraction may include a large number of patients without symptoms. *(14, 15)*.

To discriminate between these possibilities, additional catchments of different sizes should be tested. By integrating data on the presence or absence of viral particles across catchments of varying size within the same geographic region, it may be possible to estimate disease prevalence independent from knowing the average viral titer in infected stool. For example, if disease prevalence is 1 in 10,000, then approximately 50% of catchment areas representing ∼7,000 individuals would be positive (although cases may cluster within households, so additional experiments or modeling would be needed to derive precise numbers). These experiments require sampling upstream areas in the sewage system, using specialized equipment to capture time-integrated samples, and are currently underway. Once these experiments are completed in a small number of regions, they could be used to estimate disease prevalence in other regions using wastewater treatment facility data alone.

## Conclusions

These data demonstrate the feasibility of measuring SARS-CoV-2 in wastewater. The implications of this research are that WBE can be leveraged to detect population level prevalence of SARS-CoV-2 in cities across the world. In a setting where in-person testing may not be available, longitudinal analysis of wastewater can provide population-level estimates of the burden of SARS-CoV-2.

These data may help inform decisions surrounding the advancement or scale-back of social distancing and quarantine efforts based on wastewater catchment-level estimations of prevalence. Additionally, wastewater collection at the municipal or community level may allow for more granular detection of SARS-CoV-2 in cities with lower COVID-19 disease burden, thereby functioning as an early warning system to help preemptively enact public health measures prior to the widespread onset of disease.

## Methods

24 hour composite samples of raw sewage were taken from the wastewater treatment facility influent, and pasteurized at 60C for 90 min to inactivate virus. Raw sewage was then filtered through a 0.2 um membrane (Millipore Sigma) to remove bacterial cells and debris. Filters were discarded, as initial tests revealed little to no viral RNA on filters. 4 g of Polyethylene glycol 8000 (8% w/v, Millipore Sigma) and 0.9 g NaCl (0.3 M, Millipore Sigma) was then added to 40 mL filtrate, and centrifuged at 12,000 X g for 2 hours or until a pellet was visible. The viral pellet was then resuspended in Trizol (Thermofisher) for RNA extraction, followed by reverse transcription (reverse transcriptase, NEB) and qPCR (TaqMan fast advanced master mix, Thermofisher) with CDC primers (IDT) for the nucleocapsid *N* gene. Positive control (IDT) is a plasmid containing complete nucleocapsid gene from SARS-CoV-2, and used to create the standard curves for N1, N2, and N3 primers.

## Data Availability

Detailed methods available upon request.

## ACKNOWLEDGMENTS

We thank the wastewater treatment facility who worked with us in providing the samples for analysis, Penny Chisholm (MIT) and Allison Coe (MIT) for access to equipment and other supplies, Feng Zhang (Broad Institute) for reagents, Mathilde Poyet for logistical support, Katelyn Foppe for sample transportation, and Marc Lipsitch (HSPH), Karina Gin (NUS), Lee Ching Ng (NEA), and Stefan Wuertz (NTU) for helpful discussion. WPH was supported by the National Institute of General Medical Sciences. This work was supported by the Center for Microbiome Informatics and Therapeutics and Intra-CREATE Thematic Grant (Cities) grant NRF2019-THE001-0003a to JT and EJA.

